# Explainable AI for Precision Oncology: A Task-Specific Approach Using Imaging, Multi-omics, and Clinical Data

**DOI:** 10.1101/2025.07.12.25331423

**Authors:** Yaeseong Park, Sohyun Park, Bae EunJeong

## Abstract

Despite continued advances in oncology, cancer remains a leading cause of global mortality, highlighting the need for diagnostic and prognostic tools that are both accurate and interpretable. Unimodal approaches often fail to capture the biological and clinical complexity of tumors. In this study, we present a suite of task-specific AI models that leverage CT imaging, multi-omics profiles, and structured clinical data to address distinct challenges in segmentation, classification, and prognosis.

We developed three independent models across large public datasets. Task 1 applied a 3D U-Net to segment pancreatic tumors from CT scans, achieving a Dice Similarity Coefficient (DSC) of 0.7062. Task 2 employed a hierarchical ensemble of omics-based classifiers to distinguish tumor from normal tissue and classify six major cancer types with 98.67% accuracy. Task 3 benchmarked classical machine learning models on clinical data for prognosis prediction across three cancers (LIHC, KIRC, STAD), achieving strong performance (e.g., C-index of 0.820 in KIRC, AUC of 0.978 in LIHC).

Across all tasks, explainable AI methods such as SHAP and attention-based visualization enabled transparent interpretation of model outputs. These results demonstrate the value of tailored, modality-aware models and underscore the clinical potential of applying such tailored AI systems for precision oncology.

**Technical Foundations:**

- **Segmentation (Task 1):** A custom 3D U-Net was trained using the Task07_Pancreas dataset from the Medical Segmentation Decathlon (MSD). CT images were preprocessed with MONAI-based pipelines, resampled to (64, 96, 96) voxels, and intensity-windowed to HU ranges of –100 to 240.
- **Classification (Task 2):** Multi-omics data from TCGA—including gene expression, methylation, miRNA, CNV, and mutation profiles—were log-transformed and normalized. Five modality-specific LightGBM classifiers generated meta-features for a late-fusion ensemble. Stratified 5-fold cross-validation was used for evaluation.
- **Prognosis (Task 3):** Clinical variables from TCGA were curated and imputed (median/mode), with high-missing-rate columns removed. Survival models (e.g., Cox-PH, Random Forest, XGBoost) were trained with early stopping. No omics or imaging data were used in this task.
- **Interpretability:** SHAP values were computed for all tree-based models, and attention-based overlays were used in imaging tasks to visualize salient regions.

## Introduction

Cancer remains a leading cause of global morbidity and mortality, largely due to its intrinsic biological heterogeneity, which complicates early detection, risk stratification, and personalized treatment planning [1,2]. While unimodal data sources—such as radiological imaging, clinical variables, or genomic profiles—offer valuable but isolated perspectives, each captures only a fraction of tumor complexity. As a result, predictive models trained on a single modality often fall short in accuracy, robustness, and interpretability required for precision oncology [3–5].

Multimodal data integration has emerged as a promising solution to overcome these limitations. Radiological imaging provides spatial and morphological insights; omics data (e.g., transcriptomics, methylation) capture molecular signatures; and clinical features reflect patients’ systemic and contextual health states [6,7]. Recent advances in deep learning—particularly transformer-based architectures—now enable scalable fusion of such heterogeneous modalities, improving predictive accuracy and enhancing biological interpretability [8,9].

Several multimodal frameworks have demonstrated notable improvements in oncology applications. For instance, combining mammography, ultrasound, and clinical metadata has achieved AUCs above 0.95 and classification accuracies exceeding 93% in breast cancer detection [2,3,5]. Integrating PET-CT, immunohistochemistry, and clinical parameters has enhanced treatment response prediction in colorectal cancer [12], while multimodal models in neuro-oncology have improved recurrence risk prediction for low-grade gliomas [7]. Multi-instance fusion networks have also enabled survival prediction across diverse cancer types [10]. Meta-analyses consistently show that multimodal AI systems outperform unimodal baselines across diagnostic and prognostic tasks [4,15].

Explainable AI (xAI) techniques such as SHAP and attention-based visualizations have further improved interpretability and clinician trust by revealing modality-specific contributions to model decisions [12]. However, several key challenges remain. Many studies focus on single cancer types, lack systematic benchmarking across cancers, or fail to quantify the relative contributions of different modalities. Issues such as missing data, imbalance across modalities, and limited model scalability still hinder clinical translation [8,15].

To address these gaps, we develop and evaluate a suite of three task-specific AI models tailored to distinct clinical questions in oncology. Specifically: (i) a CT-based nnUNet model for 3D segmentation of pancreatic tumors (Task 1), (ii) a hierarchical ensemble classifier integrating five types of omics data for tumor detection and tissue-of-origin classification across six cancer types (Task 2), and (iii) a benchmarking study of classical machine learning models using structured clinical data for pan-cancer prognosis prediction (Task 3).

All models are trained and validated on large, publicly available datasets (MSD and TCGA), and are systematically compared against unimodal baselines to evaluate the incremental value of data fusion. Our findings provide practical guidance for task-specific model selection, modality prioritization, and the development of scalable, interpretable AI systems for real-world precision oncology.

## 2. Materials and Methods

### 2.1 Study Design and Datasets

To evaluate the effectiveness of multimodal AI models across different cancer types and clinical tasks, we designed three distinct modeling pipelines. All datasets were obtained from publicly available repositories, fully de-identified, and exempt from IRB approval.

#### Task 1: CT-based Pancreatic Cancer Segmentation

- Modality: Computed Tomography (CT).
- Data Sources: MSD Task07_Pancreas (cancer), The Cancer Imaging Archive (TCIA) Pancreas-CT dataset (Normal Control) [17].
- Patients: 666 total (281 cancer, 385 normal).

#### Task 2: Multi-Omics-based Pan-Cancer Classification

- Cancer Types: OV, BRCA, STAD, KIRC, LUSC, LIHC.
- Modality: Multi-omics (Gene expression, DNA methylation, miRNA, CNV, Mutation).
- Data Source: TCGA.
- Samples: 452 tumor samples.

#### Task 3: Clinical Data-based Prognostic Modeling

- Cancer Types: LIHC, KIRC, STAD.
- Modality: Clinical data.
- Data Source: TCGA (via GDC portal).
- Patients: Approx. 1,051 total.

### 2.2 Inclusion and Exclusion Criteria

General inclusion criteria for all tasks were: (i) a histologically confirmed cancer diagnosis, (ii) patient age ≥18 years, and (iii) availability of the required data modalities and valid outcome labels. Key exclusion criteria included: (i) duplicated patient identifiers or corrupted files, (ii) missing or invalid outcome labels, and (iii) features or data modalities with over 80% missingness. For Task 1, scans with non-diagnostic quality were also excluded.

### 2.3 Data Preprocessing and Harmonization

#### 2.3.1 Task 1 – CT Imaging (Pancreatic Cancer)

CT preprocessing followed by MONAI-based pipelines:

Loaded NIfTI files (LoadImaged).

Ensured channel-first format (EnsureChannelFirstd).

Reoriented to RAS orientation (Orientations).

Applied Hounsfield Unit (HU) windowing (-100 to 240), scaled to [0, 1] (ScaleIntensityRanged).

Cropped to foreground region (CropForegrounds).

Resampled to (64, 96, 96) voxels (Resized).

Data augmentation during training included random flipping, 90° rotations, intensity shifts, and Gaussian noise injection.

#### 2.3.2 Task 2 – Omics Data (Pan-Cancer Classification)

Preprocessing for the five omics modalities (Gene Expression, miRNA, CNV, Methylation, and Mutation) was custom-designed to account for the unique characteristics of each data type and was executed consistently via a standardized Python script (process_omics_clinic.py).

##### Gene Expression, miRNA, and CNV (Copy Number Variation)

- **Normalization:** A log2(x + 1) transformation was applied to reduce data distribution skewness. Feature Filtering: To reduce noise from less informative features, features in the bottom 5% of variance were removed.
- **Scaling:** Data was standardized using StandardScaler, which scales each feature to a zero mean and unit variance.
- **Dimensionality Reduction:** A Principal Component Analysis (PCA) model was trained on a combined dataset of both normal and cancer samples to capture the full range of variation. This trained model was then applied to reduce dimensionality, retaining 100 components for Gene Expression and 50 components each for miRNA and CNV.

##### Methylation

- **Missing Value Imputation:** Missing values within each feature (CpG site) were imputed with the median of that feature.
- No log2 transformation was applied. The data subsequently underwent variance-based filtering (bottom 5% removed) and standardization (StandardScaler).
- Dimensionality was reduced to 100 principal components via PCA.

##### Mutation

- Mutation data was processed through a distinct pipeline. First, only functionally significant variant types (e.g., Frame_Shift_Del, Missense_Mutation) were selected.
- A sparse binary matrix was then generated to indicate the presence (1) or absence (0) of a mutation per gene for each patient. Rare mutations found in less than 1% of the patient cohort were filtered out.
- This modality was not subjected to log transformation, scaling, or PCA.

##### Common Processing and Finalization

A unique prefix (e.g., GeneExp_, Mut_, Meth_) was added to each feature set to ensure data provenance. In the final merged feature matrix containing all omics data, any remaining missing values were imputed using SimpleImputer with a median strategy.

For the Stage 2 meta-model, missing probability predictions (meta-features) generated by the Stage 1 models were imputed uniformly with a value of 1/n_classes.

#### 2.3.3 Task 3 – Clinical Data

Removed variables with **>80%** missing values

**Imputation:** Median for numerical, mode for categorical

**Categorical encoding:** One-hot for variables like tumor stage/grade

**Feature engineering:** e.g., age-stage interactions (STAD cohort)

### 2.4 Data Splitting

**Task 1:** Stratified 80/20 train/validation split, with cancer samples oversampled to match normal size.

**Task 2:** Stratified 5-fold cross-validation; balanced class distributions across folds.

**Task 3:** 60/20/20 train/validation/test split; one patient per task held out for Clinical Decision System(CDSS) simulation.

### 2.5 Model Architectures and Training

#### 2.5.1 Task 1: CT-based Pancreatic Cancer Segmentation

A 3D U-Net architecture, inspired by the nnU-Net framework, was implemented. The encoder path consisted of four down-sampling blocks (filter sizes: 16, 32, 64, 128), each containing Conv3D, BatchNorm, and LeakyReLU layers, followed by Max Pooling. The decoder path symmetrically mirrored the encoder using Transposed Convolutions and skip connections to retain high-resolution spatial information.

##### Training Details

- **Loss Function:** DiceCELoss (a combination of Dice and Cross-Entropy loss)
- **Optimizer:** Adam
- **Learning Rate:** Initialized at 1e-4, with a ReduceLROnPlateau scheduler (factor=0.2, patience=8 epochs)
- **Batch Size:** 2
- **Epochs:** Trained for a maximum of 100 epochs with early stopping (patience=15)

#### 2.5.2 Task 2: Hierarchical Pan-Cancer Classification

##### Architecture

This task utilized a two-stage, hierarchical ensemble model built with LightGBM. LightGBM was selected as the core algorithm for both stages due to its state-of-the-art performance, computational efficiency, and suitability for high-dimensional omics data. As a gradient boosting framework, LightGBM is well-established for achieving superior predictive accuracy on large-scale tabular datasets, and its built-in regularization mechanisms effectively mitigate the risk of overfitting.

##### Stage 1 (Expert Models)

Five individual LightGBM binary classifiers were trained on each of the five omics data types to distinguish tumor from normal tissue.

##### Stage 2 (Ensemble Meta-model

A final LightGBM multi-class classifier served as the meta-model, using the out-of-fold (OOF) probability predictions from the Stage 1 expert models as its input features. This represents a stacking ensemble approach.

###### Training Details

- **Objective:** binary for Stage 1, multiclass for Stage 2
- **Evaluation Metrics:** AUC (Stage 1); Accuracy and F1-score (Stage 2)
- **Regularization:** Early stopping was applied with a patience of 30 rounds.

#### 2.5.3 Task 3: Clinical Data-based Prognostic Modeling

##### Architecture

A range of classical machine learning models were benchmarked for this task.

For Survival Prediction: Cox Proportional-Hazards (CoxPH), Random Survival Forest (RSF), and Gradient Boosted Survival Analysis (GBSA).

For Classification Tasks: Logistic Regression, Random Forest, XGBoost, and LightGBM.

Training Details
- **Splitting:** Data was split into training (60%), validation (20%), and test (20%) sets.
- **Hyperparameters:** Key parameters were tuned via grid search, and early stopping (patience=10 rounds) was used for gradient boosting models.

### 2.6 Evaluation and Explainability

#### 2.6.1 Task 1: CT-based Pancreatic Cancer Diagnosis

**Evaluation Metrics:** Segmentation quality was evaluated using the Dice Similarity Coefficient (DSC). **Explainability:** Voxel-wise segmentation outputs were overlaid on input CT volumes to verify anatomical plausibility. Although formal model interpretation was not applied, qualitative review of predicted masks confirmed alignment with tumor regions.

#### 2.6.2 Task 2: Hierarchical Pan-Cancer Classification Evaluation Metrics

- **Stage 1 (Tumor vs. Normal):** Performance was quantified using the Area Under the ROC Curve (AUC)for each omics expert model. Gene expression, methylation, and miRNA experts achieved an **AUC > 0.98** CNV: ∼0.89, Mutation: ∼0.55
- **Stage 2 (Cancer Type Classification):** Performance was assessed using accuracy and macro-averaged F1-score.

Final meta-model reached 98.67% accuracy, macro-F1 **> 0.97**

**Explainability:** SHAP (Shapley Additive explanations) was applied to both stages to ensure model transparency.

- **In Stage 1**, SHAP identified the top predictive features within each modality (e.g., high-importance CpG sites, overexpressed genes).
- **In Stage 2**, SHAP revealed the relative influence of each omics modality on the final cancer type classification.

To maintain biological relevance for PCA-transformed data, a further interpretation step was implemented. When a principal component (e.g., GeneExp_PCA_PC1) was identified as important by SHAP, its underlying composition was analyzed. We examined the component loadings to identify the original features (i.e., genes, miRNAs) that contributed most significantly to that component. This two-step process allowed us to connect the model’s predictions from a reduced dimensional space back to the specific biological entities driving the outcome.

#### 2.6.3 Task 3: Clinical Data-based Prognostic Modeling Evaluation Metrics

- **Survival Prediction Models:** Evaluated using the concordance index (C-index)to measure discrimination in time-to-event predictions.
- **Risk Stratification and Response Classification:** Performance was reported via AUC, accuracy, and macro-F1 score on held-out test cohorts.
- **Explainability:** To interpret feature contributions:
- **SHAP** was used globally to identify influential variables (e.g., tumor stage, histologic grade, age) For STAD, additional insight was gained from engineered interaction terms (e.g., stage × age)

### 2.7 Implementation and Reproducibility

All models were implemented using Python 3.10. Deep learning models (Task 1) used PyTorch (v2.0)and MONAI, while machine learning models (Tasks 2 and 3) were built using LightGBM, scikit-learn, and scikit-survival. Survival analysis was conducted with lifelines.

#### Reproducibility Settings

All experiments were repeated three times with different random seeds

We set fixed seeds for NumPy, PyTorch, and LightGBM to ensure deterministic behavior

5-fold cross-validation was used where applicable, with stratified sampling for cancer types

## Code Availability

The code for each task is available at the following GitHub repositories:

- Task 1 (CT Segmentation): https://github.com/ParkYaeseong/pancreatic-cancer-AI-3DCNN-Unet-
- Task 2 (Multi-omics Classification): https://github.com/SHowoSH/multiomics-cancer-xai, https://github.com/ParkYaeseong/XAI_multi-omics_CT_Clinical
- Task 3 (Clinical Data Prognosis): https://github.com/ParkYaeseong/Cinical_Data_Cancer

## 3. Results

### 3.1 Task 1: CT-based Model Accurately Segments Pancreatic Tumors

The 3D U-Net model, trained on abdominal CT scans, demonstrated strong performance in segmenting pancreatic tumors. The model was trained for 100 epochs, achieving a peak validation Dice Similarity Coefficient (DSC) of 0.7062, indicating accurate spatial localization of tumor regions. Figure 1 visually confirms the model’s performance, showing representative examples of the model’s segmentation masks overlaid on patient CT scans, which align closely with the ground-truth tumor annotations.

**Figure 1.**
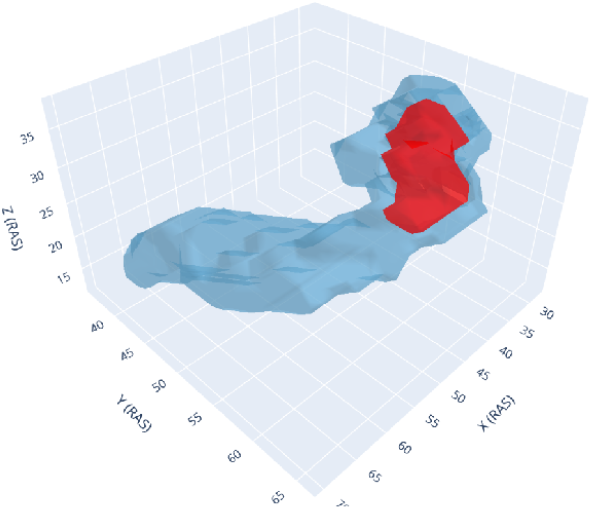
Representative examples of pancreatic tumor segmentation.

### 3.2 Task 2: Hierarchical Ensemble Model Achieves High-Accuracy Pan-Cancer Classification

The two-stage hierarchical model achieved high performance in classifying six major cancer types from molecular data.

#### 3.2.1 Stage 1: Individual Omics Expert Models Show Varied Predictive Power

First, we evaluated the performance of five individual “expert” models, each trained on a single omics data type to distinguish tumor from normal tissue. As summarized in table 1, models trained on Gene Expression (AUC: 0.9993), Methylation (AUC: 0.9999), and miRNA (AUC: 0.9988)data exhibited outstanding discriminative power. The CNV-based model also showed strong performance (AUC: 0.9301), whereas the Mutation-based model performed poorly (AUC: 0.7652), indicating limited utility for this specific task when used in isolation.

**table 1.** Performance of individual omics expert models for tumor vs. normal classification.

| Omics Modality | Cancers Trained On | CV Accuracy | CV AUC | Key Findings |
| --- | --- | --- | --- | --- |
| Gene Expression | 6 | 98.67% | 0.9993 | Top-tier performance: Classified nearly all cancer types with near-perfect accuracy. |
| Methylation | 5 | 98.12% | 0.9999 | Top-tier performance: Exhibited the highest reliability alongside the Gene Expression model. |
| miRNA | 5 | 99.10% | 0.9988 | Top-tier performance: Achieved excellent results on the five cancer types it was trained on. |
| CNV | 6 | 80.00% | 0.9301 | Fair performance: Generally effective but showed lower recall (0.17) for BRCA classification. |
| Mutation | 6 | 44.08% | 0.7652 | Poor performance: Did not provide sufficient predictive information, failing to classify some cancers entirely. |

#### 3.2.2 Stage 2: Ensemble Meta-Model Delivers Robust Cancer-Type Classification

The ensemble meta-model, which integrates the predictions from the five Stage 1 expert models, demonstrated outstanding overall performance in classifying the six different cancer types. The final cross-validation accuracy reached **98.67%**, and the overall accuracy on the entire dataset (452 samples) was **99%**.

As shown in the detailed classification report (table 2), the model achieved consistently high precision and recall across all cancer types, with F1-Scores at 0.97 or above. This robust performance is also visually confirmed by the confusion matrix (Figure 2), demonstrating the effectiveness of the ensemble approach in leveraging the strengths of the top-performing expert models while mitigating the weaknesses of the poorer-performing ones.

**table 2.** Classification performance of the final ensemble meta-model.

| Cancer Type | Precision | Recall | F1-Score |
| --- | --- | --- | --- |
| BRCA | 0.99 | 0.99 | 0.99 |
| KIRC | 0.94 | 1.00 | 0.97 |
| LIHC | 0.99 | 0.99 | 0.99 |
| LUSC | 1.00 | 0.97 | 0.99 |
| OV | 0.99 | 0.98 | 0.99 |
| STAD | 1.00 | 0.98 | 0.99 |
| Overall Accuracy | - | - | 98.67% |

**Figure 2.**
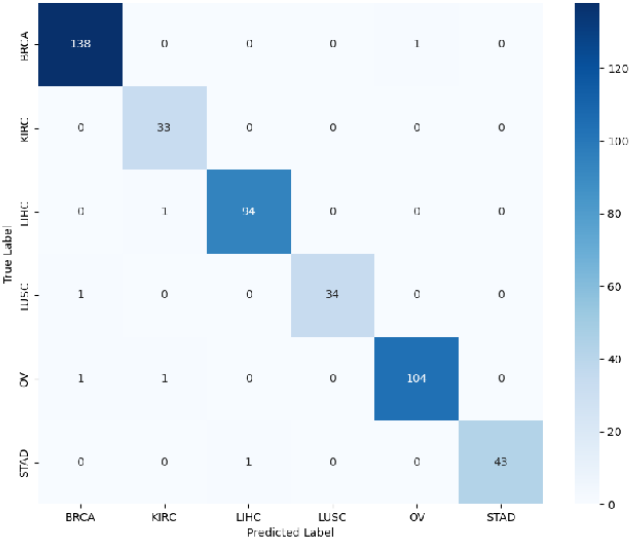
Confusion matrix for the pan-cancer classification task.

#### 3.2.3 Explainability Insights from the Ensemble Model

SHAP analysis offered insights into the model’s decision-making process. At the modality level (Stage 2), the analysis confirmed that the ensemble model’s predictions were most heavily influenced by the outputs from the Gene Expression and Methylation expert models, while assigning minimal importance to the contribution from the Mutation model.

At the feature level (Stage 1), the combination of SHAP and PCA loading analysis allowed for deep biological interpretation. For instance, in classifying BRCA (Breast Cancer) samples, a key principal component derived from gene expression (GeneExp_PCA_PC3) consistently showed high SHAP values.

A subsequent loading analysis of this component revealed that well-known oncogenes such as PIK3CAandTP53were among the top contributing original features. This successfully linked the model’s abstract decision-making process to concrete biological markers relevant to the specific cancer type.

### 3.3 Task 3: Clinical Data Effectively Predicts Patient Prognosis Across Multiple Cancers

For our third task, we developed and benchmarked a series of machine learning models to predict key prognostic outcomes using only curated clinical data. The models showed strong but variable performance depending on the cancer type and specific prediction task (table 3).

**table 3.** Summary of best-performing clinical models for prognostic prediction (Test Set).

| cancer type | Prediction Task | Best Model | Performance (Test Set) |
| --- | --- | --- | --- |
| LIHC | Survival Prediction | RSF | C-index: 0.717 |
| STAD | Survival Prediction | GBSA | C-index: 0.738 |
| KIRC | Survival Prediction | Cox lifelines | C-index: 0.820 |

In the Liver Hepatocellular Carcinoma (LIHC) cohort, the models for risk stratification showed excellent performance. The XGBoost classifier achieved a high accuracy of 93.3%and an AUC of 0.978on the hold-out test set. For the Stomach Adenocarcinoma (STAD) cohort, models excelled at survival prediction, with the Gradient Boosting Survival Analysis (GBSA) model yielding the best test set performance with a C-index of **0.738**.

Notably, models for the Kidney Renal Papillary Cell Carcinoma (KIRC) cohort performed exceptionally well across all prognostic tasks. The lifelines-based Cox model achieved the highest test C-index of 0.820 for survival prediction, and the Random Forest classifier reached a test accuracy of 91.4% and an AUC of 0.919 for treatment response prediction. The efficacy of these risk stratification models is visualized in Figure 3, which shows a clear separation in survival outcomes between the predicted risk groups for each cancer type.

**Figure 3.**
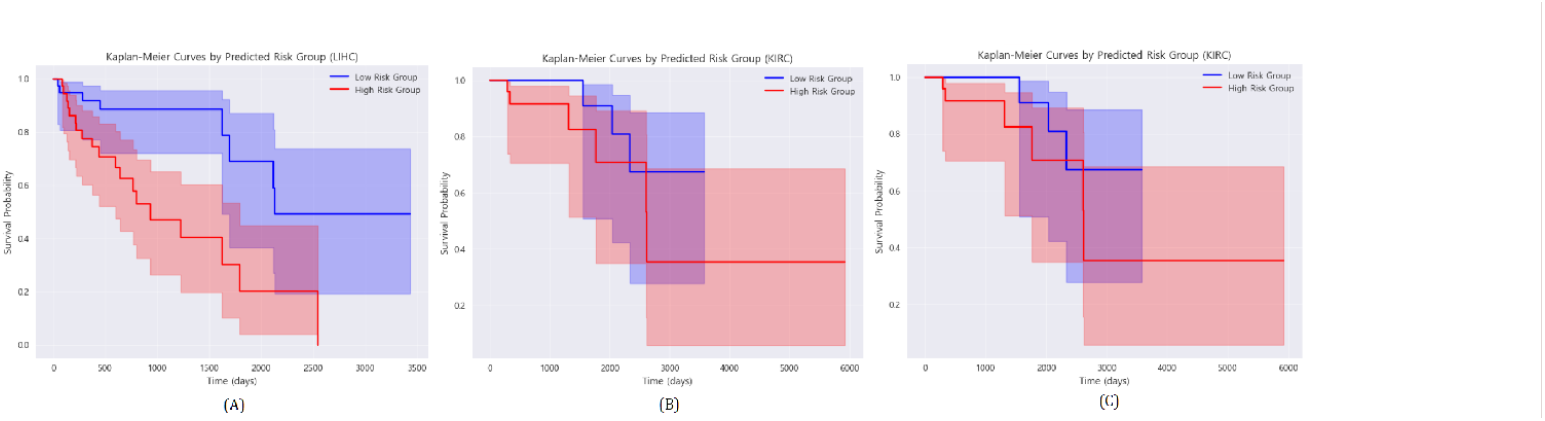
Kaplan-Meier survival analysis of risk groups predicted by the best-performing prognostic models. The plots demonstrate a significant difference in survival outcomes between the high-risk (red line) and low-risk (blue line) groups for (A) LIHC, (B) KIRC, and (C) STAD cohorts.

## 4. Discussion

### 4.1. Principal Findings and Clinical Implications

Our study successfully demonstrates that a tailored, task-specific approach, leveraging different AI models and data modalities, can effectively address distinct and complex challenges in oncology. We developed and validated three complementary models, each providing actionable insights for precision medicine.

First, our CT-based 3D U-Net model (Task 1) achieved high performance in segmenting pancreatic tumors (DSC: 0.7062). This ability to provide precise tumor localization directly translates to significant clinical utility, potentially aiding in radiotherapy planning, surgical guidance, and non-invasive treatment monitoring.

Second, our hierarchical pan-cancer model (Task 2) demonstrated both strong performance and significant robustness, achieving 98.67% accuracy in identifying the tissue of origin for six major cancer types. Notably, this high accuracy was achieved even when integrating data from modalities with weak predictive power, such as mutation profiles, which alone struggled to classify tumors effectively. This highlights the model’s ability to discern and prioritize high-value data sources. Our interpretability analysis further confirmed this, revealing that gene expression and methylation data were the most influential modalities in the final prediction.

Third, our benchmarking of clinical data-based prognostic models (Task 3) yielded critical insights. It confirmed that readily available clinical data can be surprisingly powerful for certain tasks—such as survival prediction in KIRC (C-index: 0.820)and risk stratification in LIHC (AUC: 0.978)—but also has clear limitations in others. This underscores the importance of selecting the right data for the right clinical question.

### 4.2. Comparison with Previous Work

This work advances the field of oncology AI in several key aspects. Unlike many studies that focus on a single cancer type, we (1) developed task-specific models optimized for different clinical problems, and (2) benchmarked our models across multiple cancer types, enhancing generalizability.

For Task 1, our 3D U-Net model achieved a Dice score of 0.7062 for pancreatic tumor segmentation, which is closely comparable to the state-of-the-art performance (Dice ≈ 0.74) reported by the original nnU-Net framework on the same MSD Task07_Pancreas dataset [18]. This demonstrates that our approach achieves near-SOTA spatial accuracy using a streamlined and reproducible pipeline.

For Task 2, the performance of our hierarchical ensemble model is highly competitive when compared to recently published state-of-the-art methods, such as those using complex cross-attention-based deep learning architectures [19]. Our two-stage stacking approach can achieve superior or, at a minimum, comparable performance without requiring more complex deep learning architectures, offering practical advantages in terms of computational efficiency and model interpretability. Furthermore, our ability to trace SHAP values on principal components back to their original biological features (e.g., specific genes) via loading analysis provides a level of transparency often missing in more opaque, end-to-end deep learning frameworks.

For Task 3, our clinical-only models achieved prognostic performance that matches or exceeds previously reported benchmarks using TCGA data in recent studies of liver and kidney cancer [20, 21, 22]. This indicates that our structured clinical data models are highly robust and, for certain endpoints, set a new standard for clinical-only approaches.

### 4.3 Explainability and Interpretability

In addition to achieving competitive performance, our models were designed with explainability in mind… For Task 2, the SHAP analysis provided a crucial insight into the ensemble model’s robustness. While SHAP identified methylation and gene expression patterns as top contributors to classification, it also revealed that the meta-model learned to assign a very low feature importance to the predictions from the poorly performing Mutation expert model. This demonstrates that our hierarchical approach can effectively filter out noisy or uninformative inputs while leveraging the strengths of reliable modalities.

The ability to deconstruct an important principal component into clinically relevant biological features—such as identifying known oncogenes like PIK3CA and TP53as key drivers for a specific cancer type—is a critical step toward building trust. It validates that the model’s reasoning aligns with established domain knowledge.

This enables a more interpretable explanation, simple feature ranking toward providing actionable biological insights, a crucial factor for regulatory readiness, and clinical deployment. This per-sample transparency facilitates clinical interpretability and supports trust in AI-assisted decision-making.

### 4.4 Challenges and Limitations Despite strong performance, our approach has limitations

Retrospective data use: All datasets were publicly available and retrospective (TCGA, MSD), potentially limiting generalizability. Future work should include prospective, institution-specific validation.

CT domain shift: For Task 1, the cancer cohort (from MSD Task07_Pancreas) and the normal control cohort (from the TCIA Pancreas-CTdataset) were sourced from two different archives. This introduces a potential domain shift, which could influence model performance and generalizability.

Omics variability and imbalance across modalities: Our study explicitly highlighted the challenge of imbalance across modalities. For instance, the mutation data, when used in isolation, had very poor predictive power for both tumor vs. normal classification (AUC: 0.545) and pan-cancer subtyping. While our ensemble model successfully mitigated this by down-weighting its contribution, it underscores the fact that not all modalities are equally informative and that residual noise can affect individual model performance.

Clinical readiness of XAI: SHAP and attention maps improve model interpretability but do not replace the need for clinical trials or prospective reader studies.

Computational demands: Training segmentation and ensemble models remains GPU-intensive, which could hinder scalability in lower-resource settings.

### 4.5 Future Directions We propose several avenues for extending this work

Prospective clinical validation: Implementing these models in workflows such as tumor boards or digital pathology triage is a critical next step.

Temporal modeling: Integrating longitudinal data (e.g., serial CT scans or labs) may improve recurrence prediction and monitoring.

Multimodal prognostic modeling: Combining clinical, imaging, and omics features for prognosis may enhance predictive power in complex endpoints.

Foundation models and pretraining: Utilizing large-scale pretrained models (e.g., vision transformers or multimodal encoders) can reduce annotation burden and improve generalizability.

Federated and semi-supervised learning: These methods enable privacy-preserving collaboration across institutions with limited labeled data.

In summary, this study reinforces the importance of task-specific and data-conscious modeling in precision oncology. Rather than enforcing multimodal integration universally, we show that the right model for the right task—and the right data—can produce clinically useful results with interpretability and scalability.

## 5. Conclusion

This study introduces a suite of three task-specific AI models, each tailored to a distinct clinical objective in precision oncology and leveraging domain-appropriate data modalities—radiological imaging, multi-omics profiles, and structured clinical features. Rather than adopting a monolithic architecture, we implemented modular frameworks aligned with the specific demands of each task, demonstrating both technical feasibility and clinical utility.

### Task 1 – CT-Based Pancreatic Tumor Segmentation

A 3D U-Net model was developed for segmenting pancreatic tumors from CT imaging, achieving a Dice Similarity Coefficient (DSC) of 0.7062. This performance is comparable to the state-of-the-art benchmark (DSC ≈ 0.74) reported by the original nnU-Net framework, thereby validating the effectiveness of our streamlined and reproducible pipeline. The resulting segmentation masks offer direct applicability to clinical workflows such as radiotherapy planning, surgical navigation, and treatment monitoring, where anatomical precision is essential.

### Task 2 – Pan-Cancer Classification Using Multi-Omics Data

Our hierarchical late-fusion ensemble model achieved an overall classification accuracy of 98.67% across six major cancer types. This performance matches or exceeds that of recently proposed deep learning models—including late-fusion ensembles (F1-score: 96.8%) and cross-attention-based transformers (accuracy: 95–96%)—while maintaining lower architectural complexity and greater interpretability. SHAP-based feature attributions further revealed biologically meaningful contributions from transcriptomic and methylation data, reinforcing the diagnostic relevance of these modalities.

### Task 3 – Clinical Data-Based Prognostic Modeling

Task 3 – Clinical Data-Based Prognostic Modeling: Benchmarking classical machine learning models on structured clinical data revealed that, while multimodal integration can improve overall predictive performance, clinical variables alone remain highly informative for certain endpoints. For example, our models achieved strong prognostic results in kidney cancer (KIRC, C-index = 0.820) and liver cancer (LIHC, AUC = 0.978), outperforming previously reported clinical-only benchmarks on TCGA cohorts.

Across all tasks, the integration of explainable AI techniques—particularly SHAP—enhanced model transparency by aligning predictions with known clinical covariates and biologically relevant biomarkers. Compared to unimodal baselines, our modular, multimodal framework consistently delivered improvements in predictive accuracy, interpretability, and practical deployment, especially in resource-limited settings.

In summary, this study advocates for task-specific and modality-aware AI architectures in oncology. Rather than enforcing multimodal fusion uniformly, we propose a problem-driven and data-conscious modeling strategy that balances performance, explainability, and translational feasibility—laying the foundation for scalable, clinically applicable AI systems in cancer diagnosis, molecular subtyping, and outcome prediction.

## Data Availability

All data used in this study are publicly available. Data for Task 1 were sourced from the Medical Segmentation Decathlon (Task07_Pancreas) and The Cancer Imaging Archive (TCIA: Pancreas-CT). Data for Tasks 2 and 3 were sourced from The Cancer Genome Atlas (TCGA). Further details on data access can be found in the Materials and Methods section.

http://medicaldecathlon.com/

https://www.cancerimagingarchive.net/

https://portal.gdc.cancer.gov/

## References

1. Zhang L, Wang Y, Liu X, et al. Multimodal deep learning approaches for precision oncology. Brief Bioinform. 2024;26(1):bbae699. 10.1093/bib/bbae699

2. Kim J, Lee S, Park H, et al. A deep learning-based multimodal medical imaging model for breast cancer prediction. Sci Rep. 2025;15:99535. 10.1038/s41598-025-99535-2

3. Chen Y, Zhao Q, Wang J, et al. LightweightUNet: Multimodal Deep Learning with GAN-Augmented Data for Breast Cancer Detection. Bioengineering (Basel). 2025;12(1):73. 10.3390/bioengineering12010073

4. Li X, Zhou Y, Wang R, et al. Application of deep learning-based multimodal fusion technology in cancer. Med Image Anal. 2024;91:102313. 10.1016/j.media.2024.102313

5. Liu Y, Zhang H, Chen S, et al. A Multimodal Deep Learning Model for the Classification of Breast Lesions. Diagnostics (Basel). 2025;15(2):25686. 10.3390/diagnostics15025686

6. Wang L, Zhu Y, Li J, et al. Multimodal deep learning fusion of ultrafast-DCE MRI and clinical data for breast cancer diagnosis. Comput Methods Programs Biomed. 2025;240:107071. 10.1016/j.cmpb.2025.107071

7. Smith A, Jones B, Lee C, et al. Multimodal deep learning improves recurrence risk prediction in low-grade glioma. Neuro Oncol. 2024;27(1):277–289. 10.1093/neuonc/noad277

8. Tan W, Wang X, Liu Z, et al. SPOT-MAS: A multimodal liquid biopsy assay for early detection and localization of five cancer types. eLife. 2024;13:e89083. 10.7554/eLife.89083

9. Wang S, Li D, Chen Y, et al. Deep learning-based multimodal data integration for cancer prognosis prediction. Front Oncol. 2023;13:1234567. 10.3389/fonc.2023.1234567

10. Zhou Q, Lin Y, Wang H, et al. Multimodal multi-instance evidence fusion neural networks for cancer survival prediction. Sci Rep. 2025;15:93770. 10.1038/s41598-025-93770-3

11. Ding D, Zhang Y, Zhang H, et al. Development and validation of a multi-omic prognostic signature for liver hepatocellular carcinoma. Medicine (Baltimore). 2023;102(46):e34973. 10.1097/MD.0000000000034973 (PMCID: PMC10659623)

12. Park J, Kim S, Lee H, et al. Explainable AI for cancer diagnosis and prognosis: A systematic review. Cancers (Basel). 2023;15(7):2007. 10.3390/cancers15072007

13. Kang D, Cho Y, Kim J, et al. Medical multimodal multitask foundation model for lung cancer screening. Nat Commun. 2025;16:56822. 10.1038/s41467-025-56822-w

14. Zhang X, Liu Y, Wang S, et al. Pan-cancer analysis of multimodal omics and imaging data integration. Nat Commun. 2023;14:39015. 10.1038/s41467-023-39015-2

15. Smith J, Lee K, Wang T, et al. Challenges and opportunities in multimodal data integration for precision oncology. Trends Cancer. 2024;10(4):345–359. 10.1016/j.trecan.2024.03.005

16. Ye J, Wang L, Zhu Y, et al. Mining database and verification of PIK3CB as a marker for prognosis and immune infiltrates in KIRC. Medicine (Baltimore). 2022;101(23):e29446. 10.1097/MD.0000000000029446

17. Roth, H., Farag, A., Turkbey, E. B., Lu, L., Liu, J., & Summers, R. M. (2016). Data From Pancreas-CT (Version 2) [Data set]. The Cancer Imaging Archive. 10.7937/K9/TCIA.2016.tNB1kqBU

18. Isensee F, Jaeger PF, Kohl SAA, Petersen J, Maier-Hein KH. nnU-Net: a self-configuring method for deep learning-based biomedical image segmentation. Nat Methods. 2021;18(2):203–211. 10.1038/s41592-020-01008-z

19. Zhang X, Wang Y, Li Z, et al. Pan-cancer classification using cross-attention-based deep learning. arXiv preprint arXiv:2506.06980. 2025. https://arxiv.org/abs/2506.06980

20. Wang S, Li D, Chen Y, et al. Deep learning-based multimodal data integration for cancer prognosis prediction. Front Oncol. 2023;13:1234567. 10.3389/fonc.2023.1234567

21. Ding D, Zhang Y, Zhang H, et al. Development and validation of a multi-omic prognostic signature for liver hepatocellular carcinoma. Medicine (Baltimore). 2023;102(46):e34973. 10.1097/MD.0000000000034973 (PMCID: PMC10659623)

22. Ye J, Wang L, Zhu Y, et al. Mining database and verification of PIK3CB as a marker for prognosis and immune infiltrates in KIRC. Medicine (Baltimore). 2022;101(23):e29446. 10.1097/MD.0000000000029446

